# Serum Exosomal Multi-Omic Signatures Stratify Glucose Tolerance in Cystic Fibrosis and Reveal Partial Therapeutic Reprogramming by CFTR Modulators

**DOI:** 10.1101/2025.07.07.25331056

**Authors:** Bala Umashankar, Alexander Capraro, Abhishek Vijayan, Sharon L Wong, Bibi U Nielsen, Ling Zhong, Mark Raftery, Katelin Allan, Chee Y Ooi, Sheila Sivam, Simone Visser, Bernadette J Prentice, Laura Fawcett, Zaklina Kovacevic, Lena Eliasson, Daniel Faurholt-Jepsen, Adam Jaffe, James AM Shaw, Fatemeh Vafaee, Shafagh A Waters

## Abstract

Cystic fibrosis-related diabetes (CFRD) affects up to 60% of adults with CF and contributes to poorer clinical outcomes, including accelerated lung decline and increased mortality. CFRD is often diagnosed late, with limited mechanistic insight and few tools for early detection. We profiled serum-derived exosomes from 186 individuals, 173 with CF, across two independent cohorts (Australia and Denmark), stratified by oral glucose tolerance test (OGTT) into normal (NGT), impaired (IGT), and CFRD groups. In a longitudinal subset, matched samples were collected before and after CFTR modulator therapy. Exosomes were isolated via size-exclusion chromatography and validated by NTA and TEM. Label-free proteomics and small RNA sequencing were used to profile exosomal cargo. Multi-analyte classifiers were identified using machine learning, with internal cross-validation. Exosomal profiles captured a continuum of metabolic dysfunction, with distinct signatures in CFRD including elevated PTPN1, MYO5A, and VWF (insulin resistance/hepatic dysfunction), and reduced 14-3-3ζ (β-cell dysfunction). miRNA profiles reinforced these trends, with CFRD exosomes enriched in miR-375-3p, miR-122-5p, and miR-1260a/b. CFTR modulator therapy partially reversed proteomic and transcriptomic markers of insulin resistance and hepatic dysfunction but failed to regulate β-cell-associated signatures. Machine learning models achieved high classification performance (AUC = 0.83), identifying robust multi-omic panels predictive of glucose tolerance state. This study provides the first comprehensive exosome-based multi-omics and machine learning framework for CFRD. Our findings show that serum exosomes hold promise as diagnostic and therapeutic biomarkers for early detection and monitoring of CFRD in precision CF care.

**GRAPHICAL ABSTRACT:** 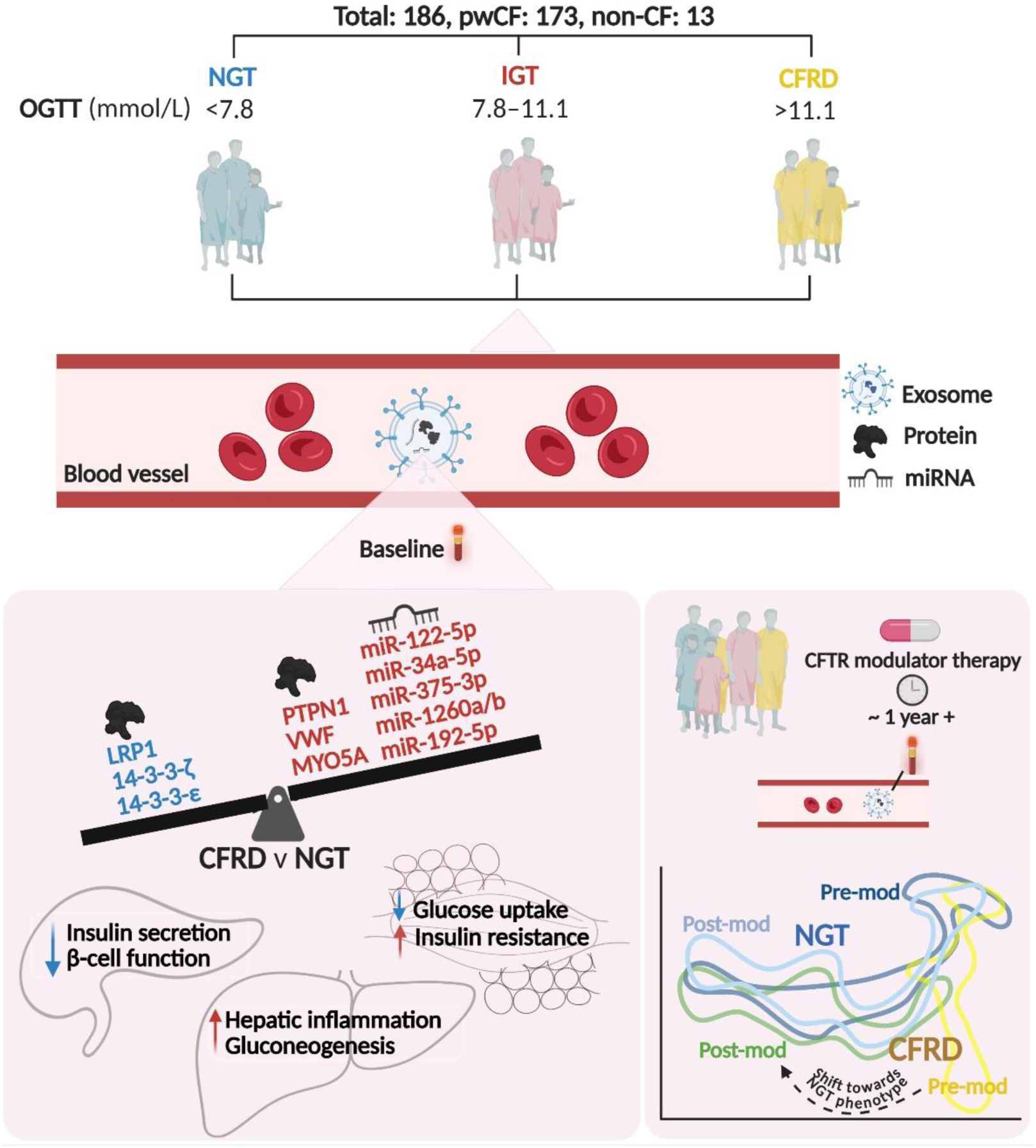

Serum-derived exosomes were isolated from pwCF and stratified into NGT, IGT, and CFRD. Exosomal signatures associated with insulin resistance, β-cell, and hepatic dysfunction were identified in CFRD compared to the NGT group. In a longitudinal follow-up cohort, exosomal signatures linked to insulin resistance and hepatic dysfunction were partially reversed in pwCFRD post CFTR modulator therapy, shifting closer toward the NGT phenotype.

## 1. INTRODUCTION

Cystic fibrosis (CF) is a multisystem disorder caused by mutations in the *CF transmembrane conductance regulator* (*CFTR*) gene, leading to impaired epithelial ion transport, chronic inflammation, and progressive multi-organ dysfunction [1]. Obstructive lung disease has been the primary cause of morbidity and mortality; however non-pulmonary manifestations are increasingly recognized as major contributors to poor long-term health outcomes and quality of life [1]. Among these, CF-related diabetes (CFRD) is one of the most common and serious comorbidities, affecting 20% of adolescents and up to 60% of adults with CF [2]. CFRD is associated with worsened nutritional status, accelerated pulmonary decline, and increased mortality [2].

CFRD pathogenesis is complex, incompletely understood and encompasses both insulin insufficiency and peripheral insulin resistance [3–7]. Contributing factors include progressive β-cell loss and function, pancreatic fibrosis, chronic inflammation, oxidative stress, and hepatic dysfunction [3–7]. Despite its clinical impact, CFRD is often diagnosed late, typically via oral glucose tolerance test (OGTT), which is limited by low adherence and poor sensitivity, particularly in early subclinical stages [8, 9]. Notably, increased pulmonary inflammation and glucose abnormalities can precede CFRD diagnosis by several years [10–12], underscoring the need for early detection of individuals at risk to mitigate long-term metabolic and respiratory decline.

CFTR modulator therapies have transformed care for people with CF (pwCF), improving pulmonary function, nutritional status, and life expectancy [4, 13]. However, their impact on glucose metabolism and CFRD remains unclear. Some studies report improvements in insulin secretion and glycemic control following CFTR modulator therapies, particularly in pwCF with normal baseline glucose tolerance [reviewed in 13]. Conversely, others find no significant metabolic benefit [14, 15, 16]. These inconsistencies highlight gaps in our understanding of how CFTR restoration influences systemic metabolism, particularly in individuals with existing or emerging glucose intolerance. This has created an urgent need for non-invasive biomarkers that can: (i) detect early metabolic dysfunction, (ii) elucidate disease mechanisms, and (iii) monitor therapeutic response.

Extracellular vesicles (EVs), including exosomes (30-150 nm) and macrovesicles (100-1000 nm), are lipid-bound nanoparticles that play central roles in intercellular communication and organ cross-talk [17]. These vesicles carry molecular cargo – proteins, lipids, metabolites, and nucleic acids such as microRNAs (miRNAs) – reflective of the physiological or pathological state of their cells of origin [17]. EVs are increasingly recognized as biomarkers and mediators in CF and type 1 and 2 diabetes mellitus (T1DM/T2DM) [18–24]. Among the various EV subtypes, exosomes are particularly well-suited for biomarker discovery in CFRD [25]. Exosomes are enriched in regulatory miRNAs and intracellular proteins tied to metabolic and endocrine function, including insulin signaling, islet health, and hepatic stress responses [23, 25, 26]. Their stability in circulation and accessibility from serum or plasma make them well suited for non-invasive, serial monitoring, and biomarker discovery [27]. In T1DM, plasma-derived exosomal miRNAs have been linked to inflammation, insulin resistance, β-cell dysfunction, and end-organ complications [23]. Similarly, a pilot study in 20 pwCF showed pro-inflammatory RNA signatures in serum exosomes reflective of both systemic and organ-specific pathology [26].

Despite these promising findings, the function and diagnostic utility of serum exosomes in CFRD remains largely unexplored. Given the overlap of pathophysiological features between CFRD and other metabolic disorders, we hypothesized that serum exosomes may contain molecular signatures associated with glucose tolerance in pwCF. Such signatures could provide novel insights into CFRD pathogenesis while offering a platform for early detection, disease stratification, and therapeutic monitoring, potentially outperforming conventional single-analyte clinical metrics.

To investigate this, we profiled serum-derived exosomes from 186 individuals, 173 pwCF, across two independent cohorts (Australia, n=87; Denmark, n=99), stratified by glucose tolerance status into normal (NGT), impaired (IGT) and CFRD groups. Exosomes were isolated via size-exclusion chromatography (SEC), validated, and characterized by label-free proteomics and small RNA sequencing. In a subset of participants, we performed longitudinal profiling before and after at least 12 months of CFTR modulator therapy. Our analysis revealed distinct exosomal signatures associated with β-cell dysfunction, insulin resistance, and hepatic dysfunction in CFRD, several of which were responsive to modulator therapy. Furthermore, machine learning-based modelling identified multi-analyte exosomal markers that accurately predict glucose tolerance status, with strong concordance between multi-variate panel and univariate differential expression analysis (**Fig. 1**).

**Figure 1.**
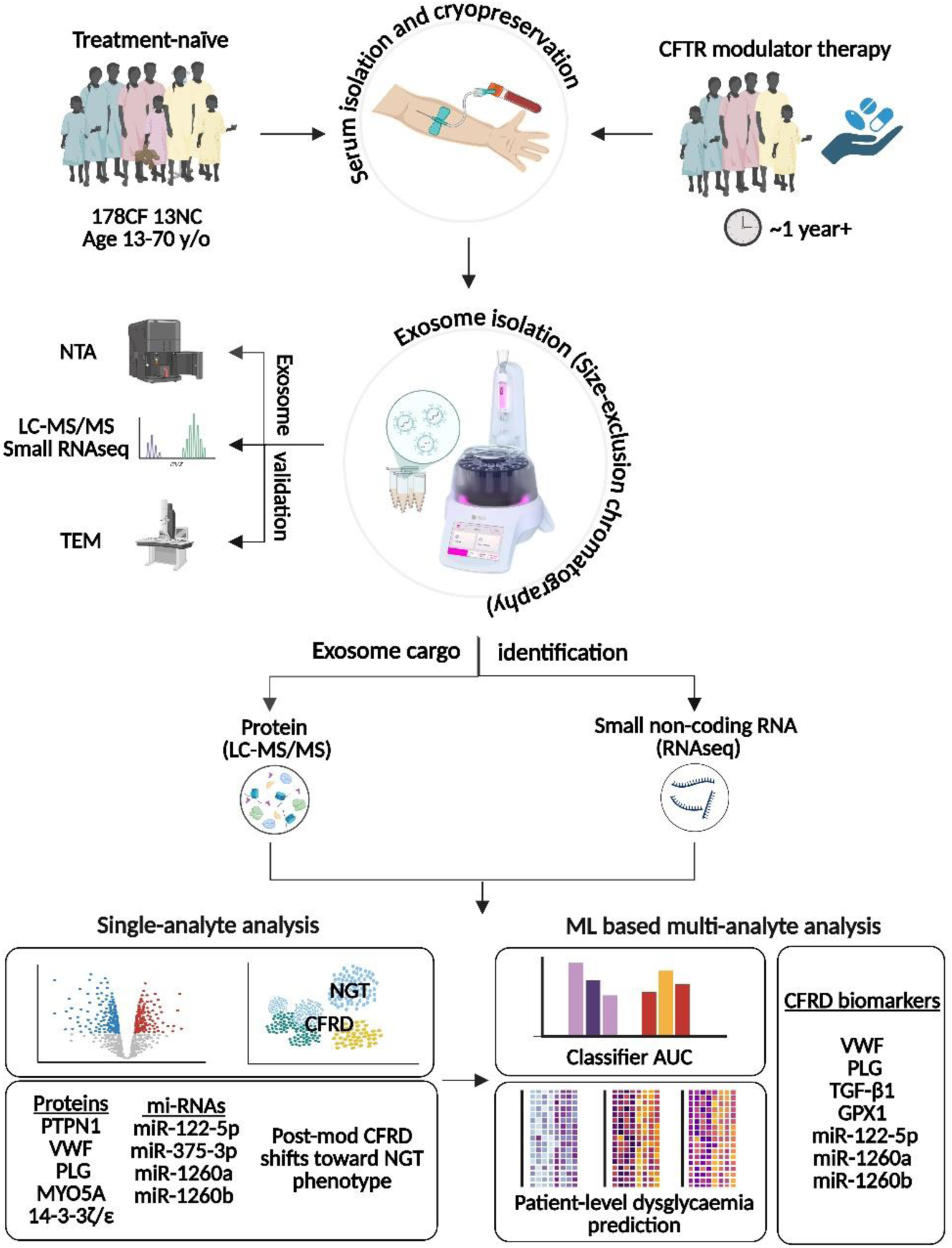
Study design and analytical pipeline for exosome-based profiling of glucose intolerance in CF. Blood samples were collected from 173 people with cystic fibrosis (CF) and 13 non-CF controls (NC) aged 13–70 years, Participants were stratified by oral glucose tolerance test (OGTT) into normal glucose tolerance (NGT), impaired glucose tolerance (IGT), and CF-related diabetes (CFRD) groups. In a subset, longitudinal samples were collected before and after at least 12 months of CFTR modulator therapy. Serum was processed by size-exclusion chromatography (SEC) to isolate exosome-enriched fractions, which were validated by nanoparticle tracking analysis (NTA) and transmission electron microscopy (TEM). Exosomal cargo was profiled using label-free mass spectrometry (LC-MS/MS) for proteomics and small RNA sequencing (RNA-seq) for miRNA analysis. Data were analyzed via univariate differential expression and multivariate machine learning (ML) models. Single analyte makers showed strong concordance with ML classifiers, which identified top exosomal biomarkers (e.g., VWF, PLG, miR-122-5p, miR-1260a/b) predictive of glucose tolerance status, with a mean AUC of 0.83 for CFRD vs NGT.

## 2. MATERIALS AND METHODS

### 2.1 Participant Recruitment and Classification

We recruited a total of 186 participants, comprising 173 pwCF and 13 non-CF controls (NC), across two independent cohorts from Australia (n=87) and Denmark (n=99). CF participants were enrolled from Copenhagen University Hospital – Rigshospitalet, Royal Prince Alfred Hospital and Sydney Children’s Hospital. Exclusion criteria included individuals with lung transplantation. NC cohorts were recruited at the University of New South Wales (UNSW), exclusion criteria included respiratory illnesses and pre-existing T1DM/T2DM.

CF participants were classified according to their clinical OGTT (two-hour blood glucose level) results into three glycaemic categories: normal glucose tolerance (NGT; <7.8 mmol/L), impaired glucose tolerance (IGT; 7.8–11.1 mmol/L), and CFRD (>11.1 mmol/L). A total of 158 pwCF had available OGTT data and were included in the classification, while 15 were excluded due to missing OGTT results.

Participants were further stratified based on CFTR modulator therapy exposure. CFTR modulator therapy included one or more of the following combinations: Tezacaftor/Ivacaftor, Lumacaftor/Ivacaftor or Elexacaftor/Tezacaftor/Ivacaftor. Amongst the pwCF cohort, 65 participants had longitudinal sampling, with matched serum samples collected before and after at least 12 months of CFTR modulator therapy. An additional 44 pwCF provided pre-modulator only samples and 49 contributed post-modulator only samples. The complete cohort distribution by health status, country, OGTT classification, and modulator therapy exposure is presented in **Table 1**.

**Table 1.** Cohort distribution and demographics. OGTT: Oral Glucose Tolerance Test, NC: non-CF Controls, CF: Cystic Fibrosis, AU: Australia, DEN: Denmark, NGT: Normal Glucose Tolerance, IGT: Impaired Glucose Tolerance, CFRD: Cystic Fibrosis-Related Diabetes, ex.: Excluded, Pre-mod: Pre-modulator, Post-mod: Post-modulator.

| Participants | Health |  | Country |  | OGTT Status |  |  |  | CFTR modulator therapy |  |  |
| --- | --- | --- | --- | --- | --- | --- | --- | --- | --- | --- | --- |
| Total | CF | NC | AU | DEN | NGT | IGT | CFRD | Unknown/ex. | Pre-mod | Pre- + Post-mod | Post-mod |
| 186 | 173 | 13 | 87 | 99 | 65 | 44 | 49 | 15 | 44 | 65 | 49 |

OGTT: Oral Glucose Tolerance Test, NC: non-CF Controls, CF: Cystic Fibrosis, AU: Australia, DEN: Denmark, NGT: Normal Glucose Tolerance, IGT: Impaired Glucose Tolerance, CFRD: Cystic Fibrosis-Related Diabetes, ex.: Excluded, Pre-mod: Pre-modulator, Post-mod: Post-modulator.

| Participants | Health |  | Country |  | OGTT Status |  |  |  | CFTR modulator therapy |  |  |
| --- | --- | --- | --- | --- | --- | --- | --- | --- | --- | --- | --- |
| Total | CF | NC | AU | DEN | NGT | IGT | CFRD | Unknown/ex. | Pre-mod | Pre- + Post-mod | Post-mod |
| 186 | 173 | 13 | 87 | 99 | 65 | 44 | 49 | 15 | 44 | 65 | 49 |

### 2.2 Serum Exosome Isolation

Blood was collected using BD Vacutainer SST II Advance Serum-gel tubes (BD Franklin Lakes, USA) and centrifuged at 3,500 × g to separate serum, which was snap frozen in liquid nitrogen and stored at −80°C. For exosome isolation, 1 mL of serum was thawed and treated with RNase A (Qiagen, Hilden, Germany). Exosomes were isolated using qEVoriginal SEC columns in the Automated Fraction Collector (IZON Science, Christchurch, New Zealand) as per manufacturer’s instructions. Serum aliquots (500uL each) were loaded and eluted with 6 mL of phosphate-buffered saline (PBS; Sigma 806552) into 12 x 1mL fractions. EV characterization was then performed in accordance with the MISEV2018 guidelines, including nanoparticle tracking analysis, transmission electron microscopy and proteomic profiling of canonical positive and negative markers [28].

### 2.3 Nanoparticle Tracking Analysis (NTA)

Fractions were diluted (1:500 in PBS) to achieve optimal particle density and analyzed on NanoSight LM10-HS (NanoSight Ltd, Amesbury, UK-Bosch Institute, University of Sydney) with a 532 nm laser. Three 30-second videos were captured per sample at 25 frames per second (fps) with default settings for minimal expected particle size, minimum track length, and blur. Data were analyzed using NTA v3.0 to determine particle size distributions and concentration.

### 2.4 Transmission Electron Microscopy (TEM)

TEM was conducted at the Electron Microscope Unit (Mark Wainwright Analytical Centre, UNSW, Sydney). Briefly, Copper TEM grids coated with formvar, and carbon (Ted Pella Inc, California, USA) were glow-discharged using a Pelco easiGlow 91000 system (Ted Pella Inc, California, USA). Purified exosome suspensions were diluted 1:1 in PBS, and 10 μL was dropped cast onto the grids. Grids were then subjected to 12 rapid water washes and negatively stained with 2% aqueous uranyl acid (UA). Excess UA was drained from the edge of the grids onto filter paper and grids were air dried. The grids were then viewed using a Technai G2 20 transmission electron microscope (FEI, Singapore) operating at 200 kV. Images were captured with a BM Eagle 2K CCD camera (FEI, Singapore).

### 2.5 Label-Free Mass Spectrometry

Proteins from fractions were lysed in lysis buffer (2% sodium deoxycholate, 50 mM Tris-HCl, pH 8.5), heated at 90°C (3 min) and pulse sonicated (5 min) on ice. Lysates were reduced (DTT, 5 mM, 37°C, 30 min) and alkylated (IAA, 10 mM, RT, 30 min) and digested with trypsin (37°C, 18 hours). Peptides were cleaned via SDB-RPS (Styrene Divinylbenzene-Reversed Phase Sulfonate, Empore, 3M) stage tips, reconstituted in 10 μL 0.1% (v/v) formic acid, and separated using nanoLC (Ultimate 3000 RSLC UPLC and autosampler system [ThermoFisher Scientific, Waltham, MA USA]) with a C18 precolumn with H_2_O:CH_3_CN (98:2, 0.1 % TFA) at 15 µL/min and a fritless nano column (75 µm x 25 cm) containing C18-AQ media (Dr Maisch, Ammerbuch-Entringen Germany). Peptides were eluted using a linear gradient of H_2_O:CH_3_CN (98:2, 0.1 % formic acid) to H_2_O:CH_3_CN (64:36, 0.1 % formic acid) at 200 nL/min over 90 minutes. Eluting peptides were ionized using positive ion nano-ESI with 2000 V to a low-volume union, and with the tip positioned ∼0.5 cm from the heated capillary (T=275°C) of a Fusion Lumos Tribrid mass spectrometer (Thermo Scientific, Bremen, Germany).

A survey scan (*m/z* 350-1750) was acquired in the orbitrap (resolution: 120,000 at *m/z* 200, target value: 3×10^6^ ions), with lockmass enabled (*m/z* 445.12003). Peptide ions (>2.5×10^4^ counts, charge states +2 to +5) were sequentially selected for MS/MS using data-dependent acquisition, maximising dependent scans within three-second cycles. Product ions were generated via collision induced dissociation (collision energy: 30; maximum injection time: 250 milliseconds; AGC target: 5×10^4^; “inject ions for all available parallelizable time” enabled) and mass analyzed in the linear ion trap. Dynamic exclusion was set to: n times =1, exclusion duration 20 seconds, ± 10ppm.

### 2.6 Database Searches and Quantification

Raw MS data were processed using MaxQuant (v2.4.2) [29], with the Andromeda algorithm [30]. Search parameters: precursor mass tolerance: ±4.5 ppm: fragment ion tolerance; ±0.5 Da; fixed modification: carbamidomethyl (C); variable modifications: oxidation (M), acetyl (N-term). Trypsin was specified as the proteolytic enzyme, allowing up to two missed cleavages. Searches used the UniProt human proteome database (2023 Release 3, June 18) [31]. Label-free quantification was performed using MaxLFQ algorithm with default parameters. Data are available at PRIDE repository [32] at https://www.ebi.ac.uk/pride/archive/projects/PXD051641 (dataset identifier PXD051641).

### 2.7 Small RNA Isolation and Sequencing

Small RNAs were isolated from exosomes using Norgen Biotek Exosomal RNA Purification Kit (Slurry Format; Norgen Biotek Corp., Ontario, Canada), and concentrated to ∼6 µL. RNA quality was assessed via the 2100 Bioanalyzer (Agilent Technologies). Libraries were prepared using QIAseq miRNA Library Kit (Qiagen, Hilden, Germany), and sequenced on the Illumina NovaSeq 6000 platform with 75bp single-end reads (Ramaciotti Centre for Genomics, UNSW Sydney, Australia). Data were processed using QIAGEN RNA Seq Portal and can be provided upon request [33].

### 2.8 Differential Expression and Pathway Analysis

Downstream analysis proteomic and transcriptomic data was performed in R [34]. Proteomic data were filtered for contaminants and low confidence proteins (< 2 unique peptides) and retained if present in >50% of the smallest group (final set of 1,550 proteins). Missing values were imputed using MissForest and data normalized by quantile transformation. Transcript data were filtered using *filterByExpr* function from the *edgeR* R package [35] to exclude lowly expressed transcripts. The remaining data were normalised using log-counts per million (log CPM). Both proteomic and transcriptomic datasets were batch-corrected using Combat batch effect correction [36], which accounts for sample origin (country) and processing batch. Differential expression was analyzed using the limma R package [37] (FDR < 0.05; FC > 1.5 for proteins, >1.2 for transcripts). Pathway enrichment was performed using QIAGEN Ingenuity Pathway Analysis (IPA; Winter Release 2023). Plots were generated in R (*ggplot* [38]) and GraphPad Prism 10.2 [39, 40].

### 2.9 Machine Learning-Based Biomarker Discovery and Predictive Modelling

The identification of diagnostic biomarker signatures was approached as a two-step process: (1) feature selection or signature panel identification, where informative features (e.g., RNAs and/or proteins) are selected, and (2) predictive modeling, in which these features were used by a classifier algorithm to stratify CF participants (i.e., CFRD vs. NGT, CFRD vs. IGT, and IGT vs. NGT) based on a training cohort. While differential analysis is a common method for feature selection, it is suboptimal for identifying features with strong predictive power [41] and lacks sensitivity to *multi-variate* signature panels, where biomarkers may only be predictive in combination. To overcome these limitations, we employed FEMPipeline, our previously developed machine learning-based biomarker discovery and predictive pipeline [42]. Proteomic and transcriptomic data from pre-modulator samples were separately pre-processed (as described above) and batch-corrected using the ComBat R package [36], accounting for country of origin. The corrected datasets were then analyzed using FEMPipeline. FEMPipeline performs six repeats of 5-fold cross-validation, yielding 30 train-test splits (each using 80% of the data as the training set and 20% held out for validation). This repetition mitigates selection bias and enhances generalizability. Within each iteration, the training data undergoes multiple feature selection methods, including uni-variate (e.g., Wilcoxon and t-tests) and multi-variate approaches (e.g., minimum redundancy maximum relevance [43], random forest recursive feature elimination [44] and Ranger [45]). Following feature selection, a panel of classifiers (L1/L2 Regularized Logistic Regression, Elastic Net, SVM with Sigmoid and Radial kernels, and Random Forest) were trained and evaluated on the held-out test data. The top-performing feature selection methods were identified based on mean area under the curve (AUC) across all iterations. Features consistently selected in ≥27 of 30 iterations were retained for further modelling. This approach ensures robustness by reducing the influence of specific training set compositions and ensuring the results are agnostic to the feature selection method used. These features were re-analyzed (excluding the feature selection step) to identify the final signature panel. This process was conducted separately for proteomic and transcriptomic data to derive distinct protein and RNA signatures.

### 2.10 Multi-omics Diagnostic Modelling

To assess the utility of a multi-omics approach, where both transcriptomic and proteomic signatures contribute to clinical outcome prediction, we applied a late fusion (decision-level integration) strategy. This approach aggregates predictions from independently trained transcriptomic and proteomic models, as previously described [46]. Specifically, we implemented integrated models using stacking ensemble [47] and delegated learning [48], combining predictions from both data types. In stacking, predictions from multiple base models (first-level classifiers) are combined by a higher-level meta-model, which learns to optimally integrate outputs. Six classification models—L1/L2 Regularized Logistic Regression, Elastic Net, SVM with Sigmoid and Radial kernels, and Random Forest—were trained separately on transcriptomic and proteomic datasets. The top two performing models from each omics were then combined using each of the six classifiers as meta-models, and stacking was performed in three configurations: i) transcriptomic-only, ii) proteomic-only, and iii) multi-omics (transcriptomic and proteomic). The highest-performing stacked models – typically those using L2 Regularized Logistic Regression – were then used to construct a delegated classifier. Delegated classification [48] dynamically selects which model to rely on based on prediction confidence. The primary model (stacked proteomic model) generates an initial prediction and confidence score. If confidence exceeds 0.7, its prediction is accepted. If not, the decision is delegated to either i) the stacked transcriptomics model or ii) the combined multi-omics stacked model. This delegated approach balances cost-effectiveness and predictive performance, allowing transcriptomic data to be measured only when needed—i.e., when the proteomic model is uncertain—thereby offering a scalable and clinically practical diagnostic strategy.

### 3. RESULTS

### 3.1 Pooling Early SEC Fractions Yields EVs Suitable for Comprehensive Proteomic and Small RNA Analysis

We optimized a size-exclusion chromatography (SEC) workflow for isolating EVs from 500 µL of human serum using qEVoriginal columns (IZON). Nano-particle rich fractions (F) were eluted between F6-F12, with F1-F5 comprising the void volume. Nanoparticle tracking analysis (NTA) confirmed that F6-F12 contained particles ranging from 100-175 nm in diameter, consistent with exosomes (n=10; **Fig. 2A**). Transmission electron microscopy (TEM) visualized vesicles with characteristic cup-shaped morphology (**Fig. 2B**).

**Figure 2.**
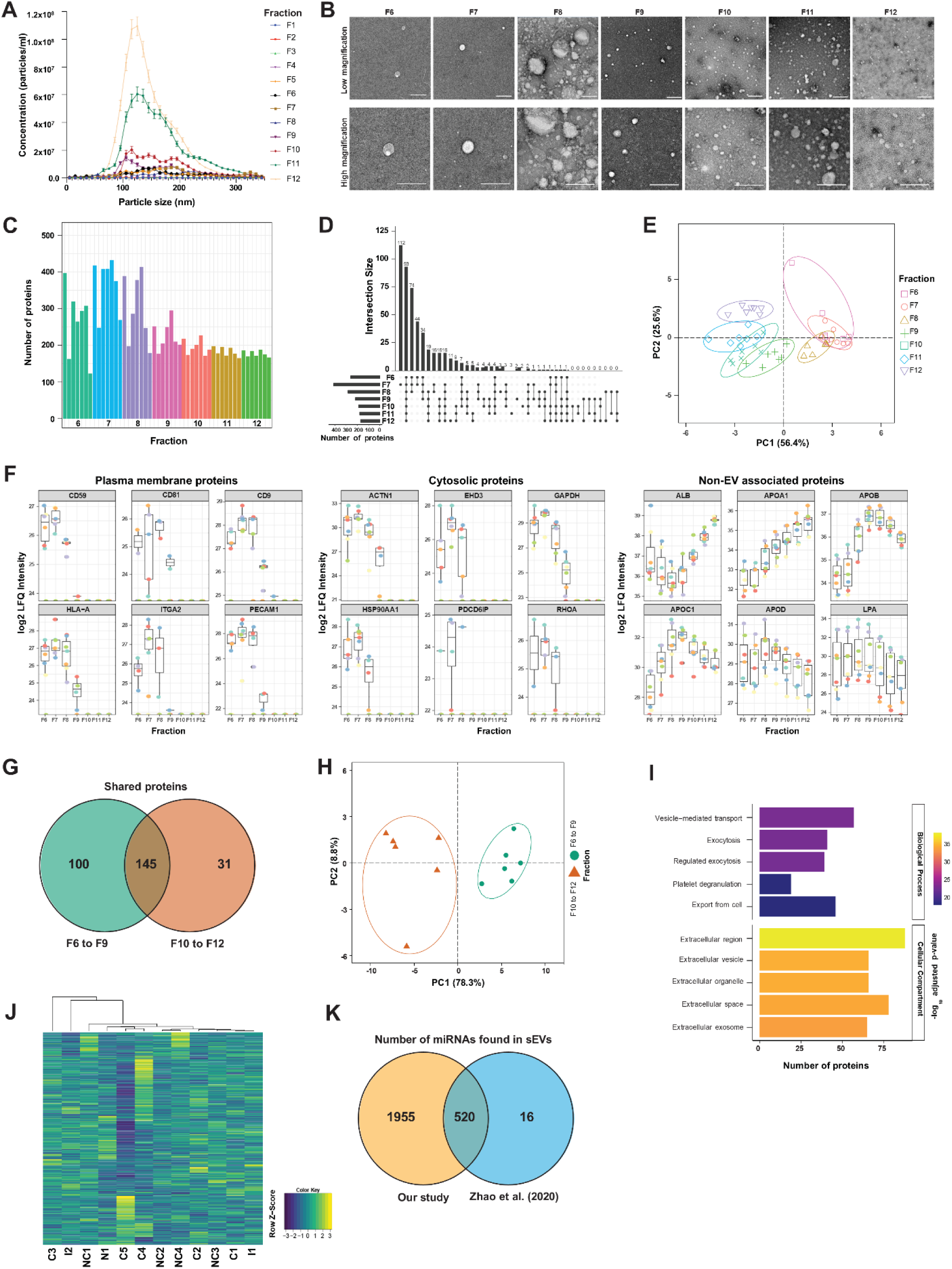
SEC-isolated early fractions yield high-purity serum EVs enriched for canonical markers and small RNAs. (A) Nanoparticle tracking analysis (NTA) of SEC fractions (F1-F12) from non-CF and CF serum (n=10) showing particle concentration and size distribution. (B) Transmission electron microscopy (TEM) of F6-F12 confirms vesicle morphology, including size uniformity and cup-shaped structure. Scale bar: 200 nm. (C) Total number of proteins identified per fraction, highlighting F7 as the most compositionally diverse. (D) Number of unique proteins per fraction, highlighting F7 as the most compositionally diverse. (E) Principal Component Analysis (PCA) of proteomic profiles distinguishes early (F6-F8) from late (F10-F12) fractions. (F) Log_2_ label-free quantification (LFQ) quantification of canonical EV markers (membrane, cytosolic), and co-isolated contaminants across fractions, box plots show mean ± SEM (n=7). (G) Venn diagram showing overlap of proteins identified in >50% of samples from pooled F6-F9 versus F10–F12. (H) PCA of pooled F6–F9 and F10–F12 proteomes confirms distinct clustering (n=6). (I) Gene Ontology (GO) enrichment of proteins unique to F6–F9 identifies strong association with vesicle transport and extracellular pathways. Bars show -log_10_ adjusted p-values. (J) Hierarchical clustering of small RNAs detected in pooled F6-F9 exosomes (n=12); color scale reflects log CPM-normalized expression. (K) Overlap of detected miRNAs with previously reported exosomal miRNAs [51].

Label-free mass spectrometry identified between 180–430 unique proteins per fraction, with F6–F8 containing the most proteins and F7 showing the greatest diversity (n=7; **Fig. 2C-D**). Principal component analysis (PCA) of proteomic profiles showed clear separation between early (F6–F8) and late (F10–F12) fractions (n=7; **Fig. 2E**). Canonical EV markers reported in Vesiclepedia [49] – including membrane proteins (CD59, CD81, CD9) and cytosolic components (ACTN1, HSP90AA1, EHD3) – were enriched in early fractions (F6-F9), while late fractions were dominated by soluble non-EV protein contaminants such as albumin and apolipoproteins [50] (**Fig. 2F**, **Table S1**). This confirms F6–F9 as exosome-enriched fractions with minimal soluble protein carryover.

To maximize yield without compromising purity, fractions F6–F9 were pooled. This pooled fraction contained 245 proteins, including 33 of the top 100 EV markers identified primarily in small EV studies [49]. In contrast, pooled fractions F10-F12 yielded only 176 proteins, with just 5 top-ranked EV markers [49] (**Fig. 2G, Table S1-2**). PCA confirmed distinct proteomic profiles between the F6-F9 and F10-12 pools (n=6; **Fig. 2H**), and pathway enrichment analysis further supported this distinction. F6-F9 proteins were enriched for vesicle-mediated transport and extracellular signaling pathways, whereas F10-F12 proteins were associated with non-vesicular immune processes (n=6; **Fig. 2I, Table S2**).

miRNA sequencing of pooled F6–F9 identified 2,475 miRNAs and 77 piRNAs with > 1 CPM (n=12, **Fig. 2J**). Of these, 520 miRNAs (21%), including six of the ten most abundant species, overlapped with previously reported exosomal miRNAs [51] (**Fig. 2K, Table S3**). Together, these data identify F6–F9 as optimal exosome-enriched serum fractions, suitable for high-yield, high-purity proteomic and small RNA profiling.

### 3.2 Serum Exosomes Reflect Progressive Insulin Resistance and Hepatic Dysfunction Across Glucose Tolerance States in CF

To identify exosomal signatures associated with glucose intolerance in CF, we stratified treatment-naive pwCF with known OGTT results (n=158) into NGT (n=65), IGT (n=44), and CFRD (n=49) groups. Proteomic profiling of serum-derived exosomes revealed progressive molecular changes across glucose tolerance states. CFRD exosomes showed increased abundance of PTPN1, MYO5A – key mediators of insulin resistance [52–57] – alongside reduced levels of insulin signalling and β-cell regulatory proteins (LRP-1, YWHA/14-3-3 family) (**Fig. 3A, Table S4**) [58–61]. Both IGT and CFRD exosomes showed increased VWF, a marker of hepatic and platelet dysfunction, and reduced abundance of coagulation-associated proteins (PLG, MYH9, FLNA, TLN1, RAP1B) [61] (**Fig. 3A, Table S3**). Notably, CTSD, a marker of hepatic inflammation and peripheral insulin resistance [63], was specifically enriched in IGT, suggesting early metabolic stress (**Fig. 3A, Table S4**).

**Figure 3.**
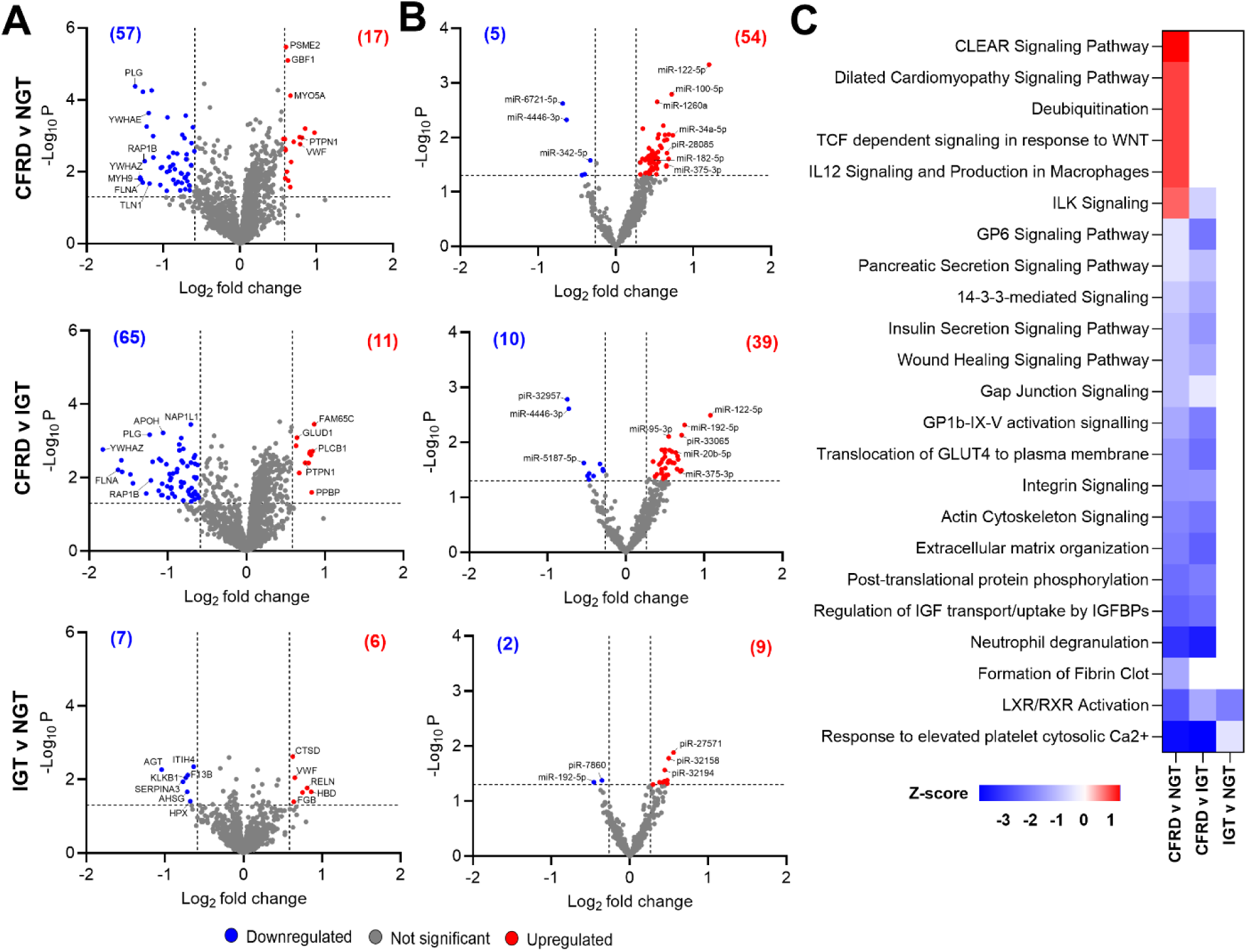
Exosomal proteomic and miRNA profiles reveal progressive metabolic and hepatic dysregulation in CFRD. (A) Volcano plots of differentially expressed exosomal proteins in CFRD v NGT, CFRD v IGT and IGT v NGT groups (treatment-naïve pwCF). Significantly upregulated (red) and downregulated (blue) proteins were defined by p ≤ 0.05, |fold change| ≥ 1.2 (B) Differentially expressed exosomal miRNAs across glucose tolerance groups, highlighting disease -associated miRNAs linked to liver dysfunction, insulin resistance, and β-cell dysfunction. (C) Heatmap of significantly enriched pathways (IPA) from proteomic data across comparisons. Red denotes positive enrichment (z-score > 0), blue denotes negative enrichment. Sample size: N (n=65), I (n=44), C (n=49).

Exosomal miRNA profiles also reflected metabolic dysfunction. CFRD exosomes exhibited an increased abundance of miR-375-3p (β-cell dysfunction) [64–66], miR-182-5p and miR-1260a (insulin resistance) [67], and miR-122-5p and miR-34a-5p (associated with CF liver disease, CFLD) [68, 69]. miR-192-5p, a marker of hepatic inflammation [70], was reduced in IGT, but increased in CFRD, indicating a trajectory of progressive hepatic dysregulation (**Fig. 3B, Table S4**).

IPA revealed distinct functional signatures in CFRD exosomes, including positive enrichment of IL-12 signalling and cardiac stress pathways (e.g. dilated cardiomyopathy), alongside suppression of insulin-signalling pathways such as IGF signalling, GLUT4 translocation, and insulin secretion (**Fig. 3C, Table S5**). Both IGT and CFRD exosomes demonstrated downregulation of LXR/RXR activation and platelet calcium signalling pathways, indicating shared early disruption of hepatic and metabolic homeostasis (**Fig. 3C, Table S5**).

Collectively, these data demonstrate that serum exosomes capture a continuum of molecular dysfunction in CF, marked by increasing inflammation, insulin resistance, β-cell and hepatic dysfunction from IGT to CFRD.

### 3.3 CFTR Modulator Therapy Partially Shifts Markers of Insulin Resistance and Hepatic Dysfunction but Not β-Cell Dysfunction in CFRD

To assess the impact of CFTR modulator treatment on systemic metabolism, we profiled serum exosomes from matched participants pre- and post-modulator therapy across glucose tolerance groups.

In CFRD, exosomes isolated from participants on CFTR modulators showed increased abundance of insulin signalling and coagulation-related proteins (LRP1, PLG and SPTA1), alongside reduced abundance of pro-inflammatory insulin resistance and mucus-associated proteins (LTF and MUC5B), indicating improved levels metabolic and hepatic profiles [52–57, 62–63] (**Fig. 4A**). In IGT, exosomes from participants on CFTR modulators showed increased abundance of insulin signaling mediator (PTPN1) and coagulation proteins (KLKB1 and F13B), but reduced abundance of β-cell survival marker (YWHAZ/14-3-3ζ) and cytoskeletal proteins (MYH9, FLNA, TLN1, RAP1B), suggesting partial metabolic remodelling with persistent β-cell vulnerability [58–62]. In NGT, changes were subtle, with elevated PTPN1 and reduced LTF and TLN1, suggesting modest metabolic remodelling (**Fig. 4A, Table S6**).

**Figure 4.**
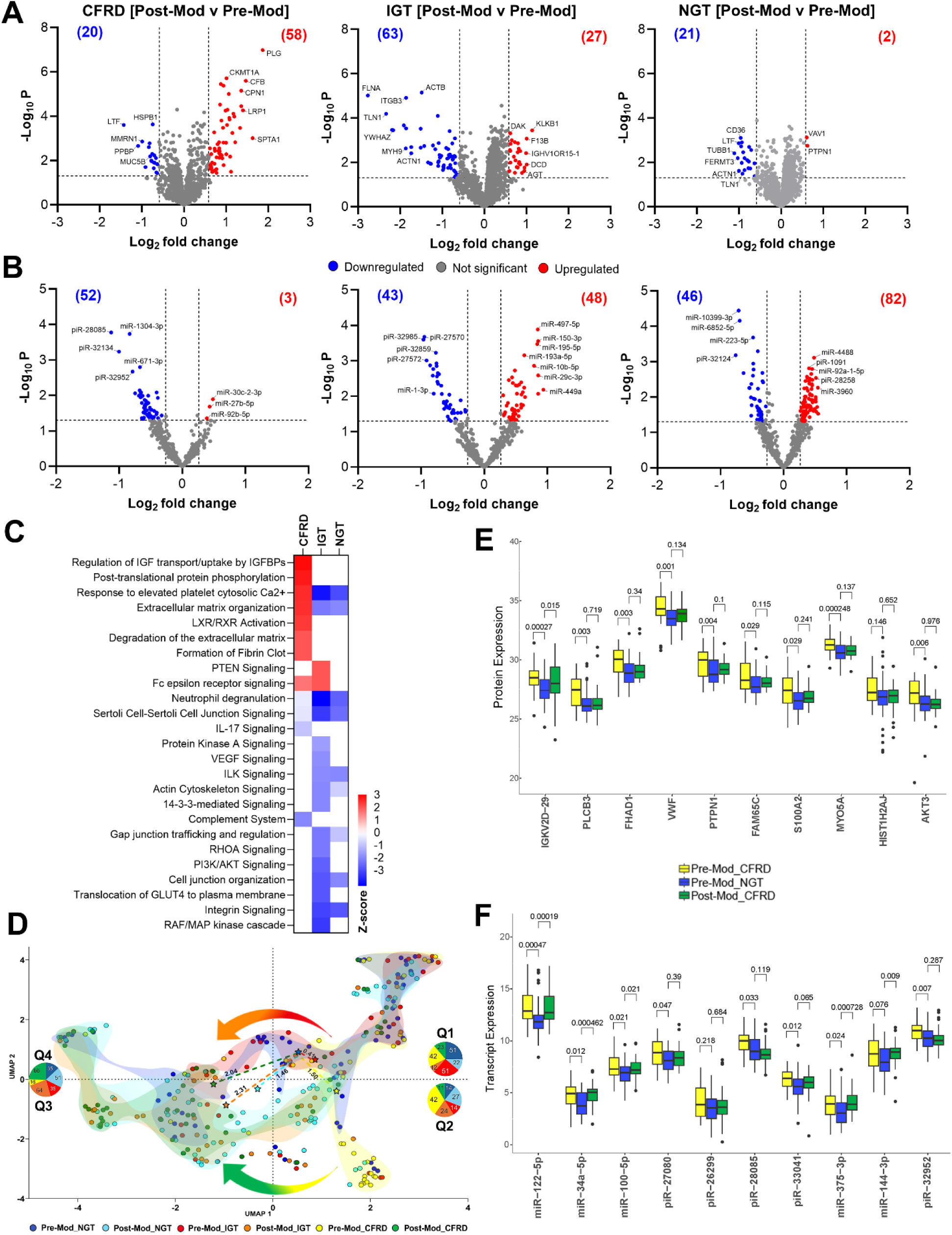
CFTR modulation partially restore insulin and hepatic exosomal signatures without rescuing β-cell function in CFRD. (A) Volcano plots of differentially expressed exosomal proteins in CFRD, IGT, and NGT groups comparing pre- and post-modulator therapy. (B) Differential expression of exosomal miRNAs across groups, highlighting changes in transcripts linked to β-cell dysfunction, insulin resistance, and liver dysfunction. Red and blue dots indicate significantly upregulated and downregulated proteins (p ≤ 0.05, |fold change| ≥ 1.2) (C) Heatmap of significantly enriched pathways (IPA) based on proteomic data. Z-scores reflect predicted activation (red) or inhibition (blue) in post-versus pre-modulator exosomes for CFRD (C, left), IGT (I, middle), NGT (N, right). (D) UMAP plot of global proteomic profiles showing post-modulator IGT, and CFRD samples shift closer to NGT samples, denoted by the trajectory lines. The colored dots and stars indicate the UMAP and centroid coordinates, respectively, for each condition. Pie charts indicate the proportion of clustering in each quadrant and the numbered dotted lines denote the Euclidean distances between pre-modulator NGT and other conditions. Top 10 differentially expressed (E) proteins and (F) miRNAs in pre-modulator CFRD vs pre-modulator NGT alongside post-modulator CFRD, ranked by log_2_ fold change. Statistical significance assessed using the Wilcoxon test with Benjamini–Hochberg correction (p ≤ 0.05). Sample size: 65 matched pre- and post-modulator, 44 pre-only and 49 post-only pwCF.

Small RNA profiling mirrored these trends. In CFRD, exosomes from participants on CFTR modulators showed increased abundance of miR-92b-5p and miR-27b-5p, both implicated in glycogen metabolism and liver disease [71–72]. In contrast, exosomes from IGT participants on CFTR modulators were enriched in miR-449a and miR-144-3p, which impair skeletal muscle insulin signalling and pancreatic β cell function, respectively [73–74] (**Fig. 4B, Table S6**).

IPA revealed that exosomes from CFRD participants on CFTR modulators gained enrichment in insulin sensitivity and liver-associated pathways (e.g. IGF transport, platelet calcium response, and LXR/RXR activation) (**Fig. 4C, Table S5)**, resembling the pre-modulator NGT profile. In contrast, exosomes from IGT participants on CFTR modulators showed enrichment of PTEN signaling and suppression of β-cell and insulin signaling pathways (14-3-3-, PI3K/AKT, and RAF/MAPK). Both IGT and NGT post-modulator exosomes showed shared negative enrichment of inflammatory and extracellular matrix remodeling pathways (**Fig. 4C, Table S5**).

To evaluate the impact of CFTR modulator therapy on circulating exosomal profiles across glucose tolerance states in CF, we applied Uniform Manifold Approximation and Projection (UMAP) embeddings derived from the proteomic data (**Fig. 4D**). Pre-modulator NGT proteome cluster in Q1 and Q3, pre-modulator IGT cluster predominantly in Q1 while pre-modulator CFRD samples are enriched in Q1 and Q2, suggesting a metabolically impaired axis (**Fig. 4D**). Post-modulator IGT and CFRD samples redistributed toward Q3/4, a region occupied by pre- and post-modulator NGT, thus indicating a therapy-associated shift of IGT and CFRD exosome cargo toward an NGT like phenotype (**Fig. 4D**). To refine this interpretation, we calculated Euclidean distances from the pre-modulator NGT centroid as a proxy for metabolic normalcy. Pre-modulator IGT and CFRD samples lay 0.41 and 1.50 units from this centroid in Q1 and Q2, respectively, while their post-treatment counterparts shifted 2.31 and 2.04 units towards Q3— suggesting reorganization rather than restoration (**Fig. 4D**). This is supported by a trajectory-like redistribution in UMAP space, where post-treatment CFRD samples form a coherent arc, moving away from the disease-associated Q2 to the stabilizing Q3 (**Fig. 4D**). Collectively, these findings reveal that modulator therapy redirects exosomal signatures along a consistent, directional path toward a distinct, regulated state rather than reverting to baseline NGT.

At a granular level, 60% of the top up- and down-regulated proteins in post-modulator CFRD overlapped with the pre-modulator NGT profile. Although the same trend was observed in miRNA, this percentage was lower (∼40%) (**Fig. 4E–F, S1**). Notably, markers of insulin resistance and hepatic dysfunction (e.g. PTPN1, VWF) were restored to NGT-like levels, while β-cell markers (YWHAZ, YWHAE, and miR-375-3p) remained unchanged. Together, these findings suggest that CFTR modulator therapy may improve systemic insulin sensitivity and hepatic function in CFRD but may not fully restore pancreatic β-cell dysfunction.

### 3.4 Machine Learning Identifies Multi-Analyte Exosomal Signatures of CFRD

To identify predictive biomarkers of concomitant dysglycemia in CF, we applied a machine learning framework to treatment-naïve serum exosome proteomic and transcriptomic datasets. Feature selection and classifier training were performed using repeated cross-validation and holdout testing to ensure robustness and generalisability (see Methods). Across 30 iterations of repeated cross-validation, Random Forest classifiers consistently outperformed alternative models. For distinguishing CFRD from NGT, proteomic features yielded a mean AUC of 0.83, while transcriptomics achieved a lower mean AUC of 0.72. IGT vs NGT proteomics performed well with a mean AUC of 0.82, while transcriptomics achieved a mean AUC of 0.59. The CFRD vs IGT comparison was best classified using the Radial Kernel SVM for proteomics (AUC = 0.82) and a logistic regression model for transcriptomics (AUC = 0.66) (**Fig. 5A, Table S7**).

**Figure 5.**
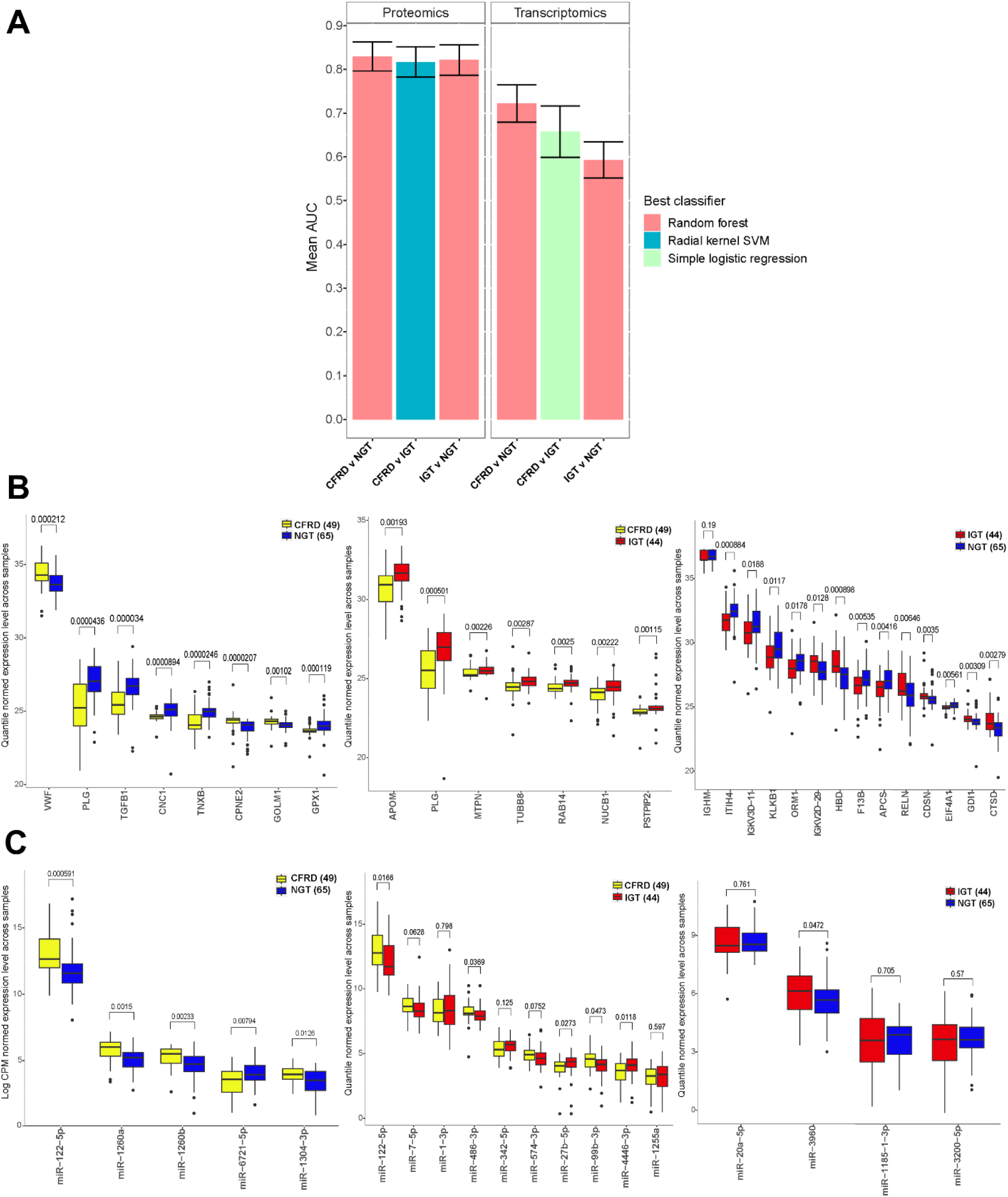
Machine learning identifies multi-analyte exosomal biomarker panels predictive of glucose intolerance in CF. (A) Mean AUC (± 95% confidence interval) for the top-performing classifiers across pairwise comparisons of CFRD v NGT, CFRD v IGT, and IGT v NGT, based on proteomics and transcriptomics data. (B) Quantile-normalised expression of top-ranked proteins across comparisons. (C) Log CPM-normalised expression of most predictive miRNAs identified for each group. Statistical comparisons were performed using the Wilcoxon test with Benjamini–Hochberg correction (p ≤ 0.05).

Multi-analyte biomarker panel indicates simultaneous modulation of multiple markers from a single analysis and the key proteins for CFRD included increased abundance of VWF, CPNE2, and GOLM1, and decreased abundance of PLG, TGFβ1, TXNB, and GPX1 (**Fig. 5B**). Among the key transcriptomic features, miR-122-5p, miR-1260a and miR-1260b emerged as top classifiers (**Fig. 5C**). These features showed strong concordance with univariate differential expression results (**Fig. 5A-B**), supporting their biological relevance and potential as integrated exosomal biomarker panels. To assess whether integrating omics layers could improve performance, we applied late fusion ensemble learning and delegated classification. The integrated multi-omics panel achieved a modest gain in CFRD vs NGT discrimination (AUC = 0.85) compared to proteomics alone but offered no significant improvement in other comparisons (**Table S8**). These results suggest that while transcriptomic data may contribute complementary information, the exosomal proteome alone provides strong diagnostic utility for glucose intolerance stratification in CF.

### 3.5 Patient-Level Predictions Demonstrate Diagnostic Potential of Exosome-Based Biomarkers

To evaluate the diagnostic utility of exosome-derived signatures, we applied the top-performing machine learning classifiers to predict glucose tolerance at the individual participant level. Using serum proteomic profiles alone, models achieved accurate stratification across OGTT-defined groups. The classifiers distinguish CFRD from NGT with 82% accuracy and 18% misclassification rate (**Fig. 6A, Table S8**). CFRD vs IGT and IGT vs NGT models achieved classification accuracies of 84% and 87%, respectively, with corresponding misclassification rates of 16% and 13% (**Fig. 6B-C, Table S8**).

**Figure 6.**
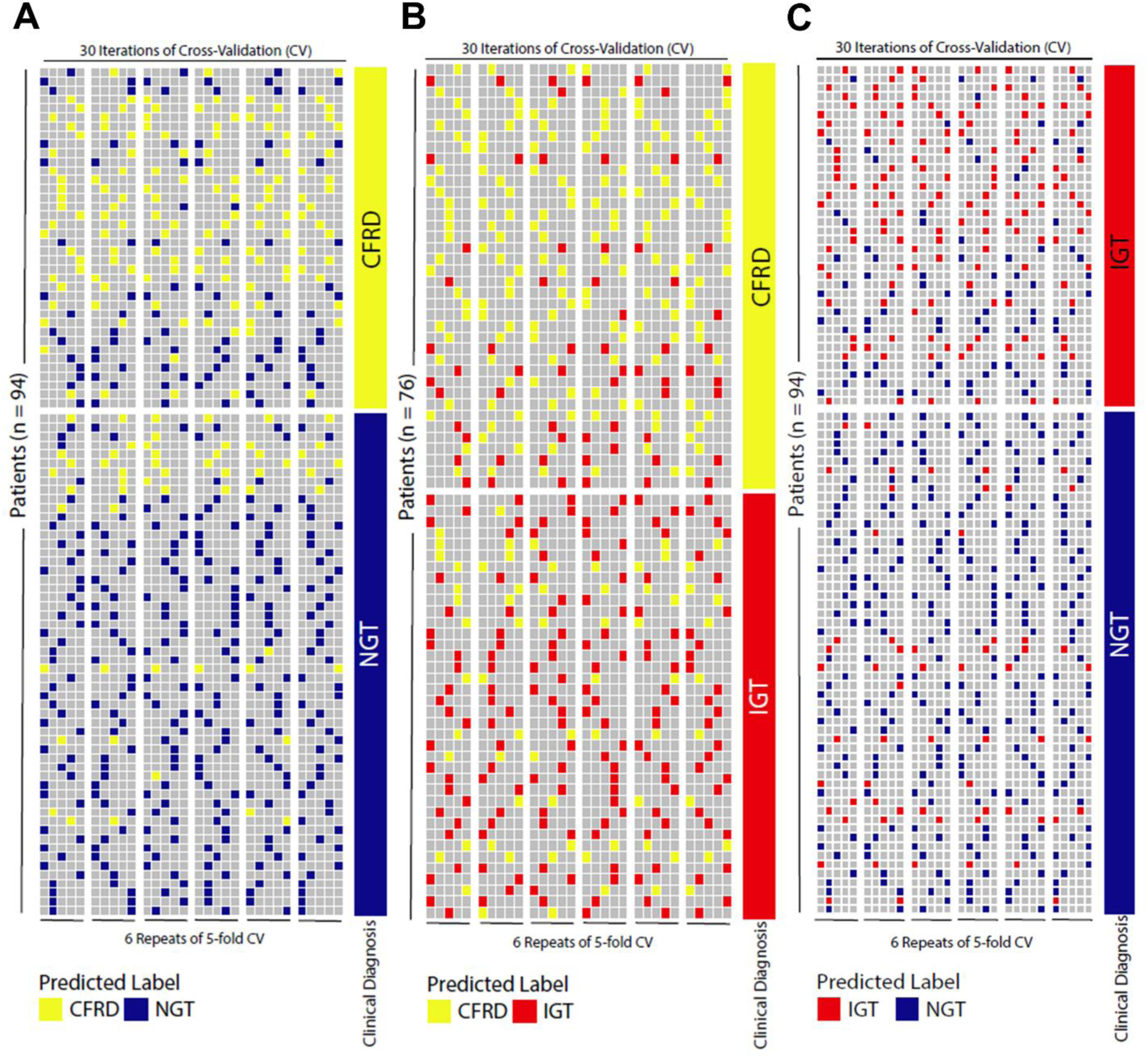
Patient-level prediction of glucose tolerance status using serum exosome proteomics aligns with clinical OGTT classification. Top-performing classifiers applied to individual-level serum proteome data accurately predicted OGGT-defined glucose tolerance status across three pairwise comparisons: CFRD vs NGT, CFRD vs IGT, and IGT vs NGT. (A-C) Heatmaps show predicted classifications across 30 iterations of 6-repeat 5-fold cross-validation (CV), with each row representing a patient and columns indicating predictions across CV runs. Clinical OGTT diagnosis is shown on the right. Sample size: CFRD (n=38), IGT (n=38), NGT (n=56).

All predictions were derived independent of clinical glucose measurements, demonstrating that exosome proteomic signatures alone can serve as minimally invasive indicators of glucose tolerance status. Notably, some misclassified individuals may represent transitional or subclinical metabolic states not fully captured by binary OGTT-defined thresholds, suggesting that EV-based profiling could uncover early or evolving metabolic dysregulation prior to clinical onset.

### 4. DISCUSSION

EVs, including exosomes and microvesicles, are gaining recognition as non-invasive biomarkers in diabetes. While urinary EVs have been explored in diabetic kidney disease [75–77], serum-derived EVs are emerging as a valuable source of biomarkers for T2DM and its complications [24, 78, 79]. In this study, we demonstrate the utility of commercial SEC columns to isolate serum exosomes for downstream applications in CF and provide evidence that glucose intolerance modulates serum exosomal protein and miRNA profiles. To our knowledge, this is the first study to apply an integrated exosome-based multi-omics and machine learning framework to stratify glucose tolerance states and assess modulator therapy response in CF. In doing so, we have generated the first proteomic and small RNA atlas of serum-derived exosomes across glucose tolerance states in CF, creating a bioresource for future biological discovery and clinical translation.

Serum-derived exosomes from CFRD participants showed proteomic signatures consistent with insulin resistance and hepatic dysfunction, including elevated levels of PTPN1, MYO5A, and VWF. In other studies, murine PTPN1 knockout models have shown improved insulin signalling and secretion [52–55]. In humans, PTPN1 and MYO5A abundance is consistently higher in insulin-resistant tissues [56], where MYO5A impairs GLUT4 translocation, glucose uptake [57] and has been reported to increase insulin release [80]. Elevated VWF in exosomes has been linked with insulin resistance and T2DM [81–83]. Conversely, CFRD exosomes have shown reduced abundance of 14-3-3 proteins (YWHAZ, YWHAE) and LRP-1. 14-3-3 proteins protect β-cells under stress, suppressing apoptotic signalling [58–60], whereas LRP-1 supports glucose uptake through GLUT4 and maintains hepatic insulin receptor expression [61]. Together, this constellation of exosomal protein changes – elevated PTPN1, MYO5A, and VWF with reduced 14-3-3 and LRP-1 – mirrors a molecular phenotype consistent with systemic insulin resistance, β-cell, and hepatic dysfunction. Pathway analysis further corroborated these findings in CFRD exosomes, indicating downregulation of insulin secretion pathways and extra-pancreatic pathways that maintain glucose homeostasis (LXR/RXR activation, translocation of GLUT4 to plasma membrane, and regulation of IGF transport and uptake) [84, 85].

CFRD exosomes also contained elevated levels of miR-375-3p and miR-1260a, both implicated in β-cell dysfunction and insulin resistance. miR-375 impairs insulin exocytosis by targeting vesicle docking machinery in pancreatic islets and is a proposed biomarker for islet damage and diabetes onset in both animal and human models [64–66]. miR-1260a, found to be upregulated in prediabetic individuals, may modulate PTPN1 expression and interfere with insulin sensitivity [67]. Their increased abundance in CFRD exosomes may not reflect tissue stress but could contribute to paracrine propagation of metabolic dysfunction.

Importantly, the impact of CFTR modulator therapy on exosomal biomarkers revealed differentia restoration across biological systems. Post modulator CFRD exosomes showed partial normalization of PTPN1, MYO5A, VWF, and LRP-1 abundance toward NGT-like levels, suggesting recovery of insulin sensitivity and improved hepatic signaling. In contrast, neither differential expression analysis nor pathway analysis provided evidence for recovery of insulin secretion or β-cell function post-modulator therapy. These findings suggest CFTR modulation may improve glucose tolerance in CFRD primarily by restoring peripheral insulin sensitivity, rather than reversing β-cell function. This aligns with clinical studies reporting improved glycemic profiles in the absence of increased insulin secretion [15, 86]. Whether the lack of β-cell recovery reflects irreversible damage or delayed recovery not yet captured by circulating exosomes remains unclear and warrant longitudinal assessment. However, low CFTR expression in β-cells may also explain this absence of response to modulator therapy [5, 87].

CFLD is a known risk factor for CFRD, and hepatic dysfunction is tightly linked to impaired glucose regulation and diabetes [6, 88]. In our study, CFRD exosomes were enriched in miR-122-5p and miR-34a-5p, also elevated in CFLD [68, 69] and T2DM with liver disease [89]. Accompanying proteomic hallmarks of hepatic dysfunction – increased VWF and decreased PLG – mirror markers of endothelial stress and impaired haemostasis in chronic liver disease [62]. While VWF and PLG levels partially normalised with CFTR modulator therapy, persistent dysregulation of liver-associated miRNAs (miR-122-5p, miR-34a-5p) and cytoskeletal proteins suggest incomplete recovery. These findings imply that hepatic benefit from CFTR modulator therapy may be delayed, incomplete, or non-cell autonomous.

Interestingly, miR-192-5p showed a biphasic profile – reduced in IGT but elevated in CFRD – consistent with its role in hepato-protection under stress-induced acute liver damage and hepatic inflammation [90, 91]. Its increase in CFRD may signal transition from adaptive to maladaptive hepatic states. miR-192-5p has also been shown to activate macrophages responses and promote IL-12 -driven hepatic inflammation [92], a pathway that was also enriched in the CFRD proteome. Together, these data support a mechanistic axis linking exosomal miRNA signalling, immune activation, and hepatic-driven insulin resistance in CFRD.

CFTR dysfunction in the hepatobiliary system leads to altered bile acid signalling, ductal obstruction, progressive liver inflammation and fibrosis [93, 94]. These alterations impair hepatic insulin clearance, elevate systemic insulin exposure and promote peripheral resistance [94]. Impaired hepatic glucose handling further compromises skeletal muscle glucose uptake, aggravating hyperglycemia [94]. The interplay between liver function and glucose regulation provides a plausible basis for CFLD as a risk factor for CFRD [6]. Our findings suggest that exosomes may serve not only as reporters but also as functional communicators of liver-pancreas crosstalk in the context of CFTR dysfunction. While serum EVs reflect this interplay, it remains possible that they mirror downstream effects rather than causal drivers. Longitudinal, mechanistic studies will be essential to define the temporal sequence and functional relevance of these interactions.

Our machine learning framework identified robust multi-analyte exosomal signatures that predicted glucose tolerance status with high internal accuracy. Proteomic classifiers consistently outperformed transcriptomic classifiers, achieving AUCs >0.8 across comparisons, whereas miRNA-based models performed more modestly. This discrepancy may reflect greater temporal variability or lower biological stability in circulating small RNAs. Interestingly, similar trends have been reported in other disease models, where exosomal proteomes exhibited strong classification power [95–97].

We also tested whether integrating omics layers could improve prediction. A late-fusion and delegated learning strategy yielded only marginal gains in AUC over proteomics alone, suggesting the exosomal proteome provides a strong independent signal. Key features of the CFRD vs NGT model – VWF, PLG, miR-122-5p, miR-1260a and miR-1260b were identified in both univariate and multi-variate analyses strengthens their biological and diagnostic relevance. These markers are each linked to liver injury, inflammation, and diabetic metabolic dysfunction [67, 98, 99].

Patient-level predictions showed strong concordance with OGTT-defined classification, suggesting their translational utility. Misclassified participants may represent early or transitional states of dysglycemia not captured by static OGTT thresholds, highlighting the potential of exosomal profiling to detect subclinical or evolving disease states. As such, serum EVs may be valuable not only for diagnosis, but also for metabolic surveillance in at-risk individuals.

For clinical translation, the biomarker panel should be validated in an independent CF cohort using multiplex targeted assays (e.g., qPCR, ELISA), with blinded classification. It will also be important to determine whether these biomarkers can be detected directly in serum or plasma without exosome isolation, enabling rapid, scalable diagnostics for routine care.

## CONCLUSION

This study establishes serum-derived exosomes as a powerful and minimally invasive platform for capturing the molecular complexity of glucose dysregulation in CF. By integrating proteomic and transcriptomic profiling with machine learning, we identified multi-analyte exosomal signatures that stratify glucose tolerance states and reflect differential responsiveness to CFTR modulator therapy. Our findings highlight the diagnostic potential of exosomal biomarkers—particularly those linked to insulin resistance, β-cell dysfunction, and hepatic stress—and demonstrate their ability to detect subclinical or transitional metabolic states. While proteomic signatures emerged as the most robust classifiers, the combined insights from multi-omics analysis offer a window into the systemic and organ-specific drivers of CFRD. These results lay the foundation for developing exosome-based diagnostic tools and personalized metabolic surveillance strategies in CF, with future efforts focused on clinical validation, longitudinal tracking, and translation into scalable assays for routine care.

## AUTHOR CONTRIBUTIONS

**Bala Umashankar:** Investigation, Data Curation, Formal Analysis, Visualization, Writing – Original Draft, Writing – Review & Editing. **Alexander Capraro:** Methodology, Investigation, Data Curation, Formal Analysis, Visualization, Writing – Original Draft, Writing – Review & Editing. **Bibi U Nielsen:** Resources, Project Administration, Data Curation, Validation, Writing – Review & Editing. **Abhishek Vijayan:** Data Curation, Formal Analysis, Visualization, Writing – Review & Editing. **Sharon L Wong:** Methodology, Data Curation, Investigation, Writing – Review & Editing. **Ling Zhong:** Methodology, Investigation, Data Curation, Resources. **Mark Raftery:** Methodology, Investigation, Data Curation, Resources. **Katelin Allan:** Data Curation, Writing – Review & Editing. **Chee Y Ooi:** Recourses, Writing – Review & Editing. **Sheila Sivam:** Data Curation, Writing – Review & Editing. **Simone Visser:** Resources, Data Curation, Writing – Review & Editing. **Bernadette J Prentice:** Data Curation, Writing – Review & Editing. **Laura Fawcett:** Data Curation, Writing – Review & Editing. **Zaklina Kovacevic:** Writing – Review & Editing. **Lena Eliasson:** Writing – Review & Editing. **Daniel Faurholt-Jepsen:** Funding Acquisition, Resources, Supervision, Project Administration, Writing – Review & Editing. **Adam Jaffe:** Resources, Writing – Review & Editing. **James AM Shaw:** Conceptualization, Funding Acquisition, Writing – Review & Editing. **Fatemeh Vafaee:** Conceptualization, Funding Acquisition, Methodology, Formal Analysis, Resources, Supervision, Writing – Review & Editing. **Shafagh A Waters:** Conceptualization, Funding Acquisition, Project Administration, Methodology, Investigation, Formal Analysis, Validation, Resources, Supervision, Writing – Original Draft, Writing – Review & Editing.

## ACKNOWLEDGEMENTS

The authors thank the study participants and their families for their contributions. The authors thank Saeideh Ebrahimkhani (miCF, UNSW) for conducting NTA on exosomal fractions. Small RNA Sequencing were conducted at the Ramaciotti Centre for Genomics at UNSW. Mass spectrometric raw data were obtained at the Bioanalytical Mass Spectrometry Facility within the Mark Wainwright Analytical Centre (UNSW). BP is supported by Sydney Children’s Hospitals Network Kids Research Clinician Researcher Fellowship funded by Sydney Children’s Hospitals Foundation. LF is supported by the Rotary Club of Sydney Cove/ Sydney Children’s Hospital Foundation and UNSW Postgraduate Award Scholarships. LE is supported by the Swedish Research Council (project 2024-03597) and Strategic Research Area Exodiab (2009-1039) and The Swedish Foundation for Strategic Research (IRC15-0067), Barndiabetesfonden. SAW is supported by Australian National Health and Medical Research Council grant (NHMRC_APP1188987) and UNSW Scientia award.

## ETHICS APPROVAL AND PATIENT CONSENT STATEMENT

Ethics approval was granted by the Sydney Children’s Hospital Network (HREC/16/SCHN/120) and Danish Ethical Committee (H-16022305/H19085530) and written informed consent was obtained from all participants.

## FUNDING STATEMENT

This work was supported in part by an Australian National Health and Medical Research Council grant (NHMRC_APP1188987), CFRD-SRC-019 Grant (CF Trust), and Vertex Innovation Grant (2018), Sydney Children’s Hospitals Foundation.

## DATA AVAILABILITY STATEMENT

The data that support the findings of this study are available on request from the corresponding author.

## CONFLICT OF INTEREST DISCLOSURE

The authors declare no conflicts of interest.

## Notes

### Competing Interest Statement

The authors have declared no competing interest.

### Author Declarations

Ethics approval was granted by the Sydney Childrens Hospital Network (HREC16-SCHN-120) and Danish Ethical Committee (H-16022305-H19085530) and written informed consent was obtained from all participants.

